# Longitudinal progression of blood biomarkers reveals a key role of astrocyte reactivity in preclinical Alzheimer’s disease

**DOI:** 10.1101/2024.01.25.24301779

**Authors:** VR Varma, Y An, PR Kac, M Bilgel, A Moghekar, T Loeffler, D Amschl, J Troncoso, K Blennow, H Zetterberg, NJ Ashton, SM Resnick, M Thambisetty

## Abstract

Defining the progression of blood biomarkers of Alzheimer’s disease (AD) is essential for targeting treatments in patients most likely to benefit from early intervention. We delineated the temporal ordering of blood biomarkers a decade prior to the onset of AD symptoms in participants in the Baltimore Longitudinal Study of Aging. We show that increased astrocyte reactivity, assessed by elevated glial fibrillary acidic protein (GFAP) levels is an early event in the progression of blood biomarker changes in preclinical AD. In AD-converters who are initially cognitively unimpaired (N=158, 377 serial plasma samples), higher plasma GFAP levels are observed as early as 10-years prior to the onset of cognitive impairment due to incident AD compared to individuals who remain cognitively unimpaired (CU, N=160, 379 serial plasma samples). Plasma GFAP levels in AD-converters remain elevated 5-years prior to and coincident with the onset of cognitive impairment due to AD. In participants with neuropathologically confirmed AD, plasma GFAP levels are elevated relative to cognitively normal individuals and intermediate in those who remain cognitively unimpaired despite significant AD pathology (asymptomatic AD). Higher plasma GFAP levels at death are associated with greater severity of both neuritic plaques and neurofibrillary tangles. In the 5XFAD transgenic model of AD, we observed greater GFAP levels in the cortex and hippocampus of transgenic mice relative to wild-type prior to the development of cognitive impairment. Reactive astrocytosis, an established biological response to neuronal injury, may be an early initiator of AD pathogenesis and a promising therapeutic target.

## Introduction

Blood biomarkers are on the verge of use in routine clinical practice to aid in diagnosis of Alzheimer’s disease (AD) in patients with clinical symptoms of cognitive impairment (1, 2). Furthermore, their utility in defining the earliest stages of the disease in individuals even without manifest cognitive impairment is the rationale for newly proposed biological criteria for AD as well as for emerging direct-to-consumer blood tests to predict an individual’s risk of AD (3, 4). Blood biomarkers of AD are also increasingly relevant in screening individuals for recruitment into clinical trials to identify those likely to show neuropathological features of AD (5). With the US Food and Drug Administration’s (FDA’s) recent approval of monoclonal antibodies targeting brain amyloid deposition as novel treatments in symptomatic AD (6), there is growing interest in testing such emerging treatments in preclinical AD, before the onset of overt cognitive impairment and functional decline (7).

Identifying blood biomarkers at the earliest stages of AD progression in asymptomatic individuals is an important prerequisite to targeting emerging, experimental, disease-modifying treatments in those most likely to benefit from early intervention. In this context, it is critical to define the precise temporal progression of blood biomarkers of AD that may reflect distinct, sequential molecular features of disease progression. Furthermore, such biomarkers of preclinical AD may also hold promise as surrogate markers of disease modification by novel experimental treatments if they reflect biologically relevant characteristics of AD pathogenesis and/or progression.

The most extensively studied blood biomarkers in AD are those that reflect brain beta-amyloid (Aβ42 and Aβ40) deposition, accumulation of phosphorylated tau (p-tau), neurodegeneration (neurofilament-light chain; NfL) and astrocyte reactivity (glial fibrillary acidic protein; GFAP) (8–10). Most studies testing their association with AD risk and progression have been cross-sectional analyses or performed in cohorts with limited longitudinal follow up (5, 10–13). We recently reported on longitudinal plasma biomarker trajectories relative to brain amyloid changes in cognitively normal older individuals (14). However, few studies have tested the association of these blood biomarkers with measures of AD neuropathology quantified by postmortem examination of the brain (13, 15, 16) which remains the gold standard for confirmation of a clinical diagnosis of AD.

In the present study, our primary goal was to delineate the temporal ordering of blood biomarkers a decade prior to the onset of AD symptoms in participants in the Baltimore Longitudinal Study of Aging (BLSA). We also examined the association of these biomarkers with severity of AD neuropathology quantified by postmortem assessment of the brain as well as with clinical symptoms in individuals with autopsy-confirmed diagnosis of AD. Based on the results of these primary analyses, our secondary aims were to test whether plasma concentrations of GFAP and other blood biomarkers differed by *APOE ε*4 status and to test associations between brain and plasma concentrations of GFAP in the 5XFAD transgenic mouse model of AD.

## Methods

### Participants: AD converters – cognitively unimpaired

AD converters (n = 158) and cognitively unimpaired participants (CU, n = 160) were from the Baltimore Longitudinal Study of Aging (BLSA), a prospective cohort study that began in 1958 and is administered by the National Institute on Aging (NIA) (17, 18).

AD converters included participants with mild cognitive impairment (MCI) due to AD (n=47) or dementia due to AD (n=111) with plasma samples collected at three timepoints (+/− 2 years) i.e. at symptom onset, 5 years prior to symptom onset and 10 years prior to symptom onset. CU remained cognitively unimpaired throughout the entire period of plasma sample collection. CU were sex, race, age and visit year-matched to AD converters at all three timepoints. The total sample included 318 participants with 756 visits.

### Participants: postmortem sample

Postmortem participants in the autopsy program of the BLSA have been described in detail previously (19). Three groups of participants were included in the current analyses: Alzheimer’s disease (AD; n=28); cognitively normal (CN; n=19) and asymptomatic AD (ASY; n=18). AD participants had clinical symptoms (i.e., cognitive impairment) of AD during life assessed by longitudinal cognitive assessments at consensus diagnosis conferences (20) with confirmation of AD pathology at autopsy as determined by an expert neuropathologist (21). CN participants were cognitively unimpaired during life and without significant AD pathology at autopsy. ASY participants have been described extensively previously (22). These participants were cognitively unimpaired during life as assessed by longitudinal neuropsychological assessments but had significant AD neuropathology at autopsy including both neuritic plaques and neurofibrillary tangles. Similar to AD subjects, all ASY participants had CERAD scores >1 (either 2 or 3) while all CN participants had CERAD scores ≤1 (either 0 or 1) (22).

Plasma specimens from postmortem participants were collected approximately 4 years prior to death.

The BLSA study protocol has ongoing approval from the Institutional Review Board of, National Institutes of Health. Written informed consent was obtained at each visit from all participants.

### APOE genotyping

Participants were classified as either *APOE ε*4+, a carrier of one or two *ε*4 alleles, or *APOE ε*4-, a non-carrier of the *ε*4 allele. *APOE* genotype was performed either by polymerase chain reaction (PCR) amplification wth restriction isotoping as described previously (23), or the TaqMan method (24).

### Animals

All animal experiments and procedures conformed to the Austrian guidelines for the care and use of laboratory animals (Tierversuchsgesetz 2012-TVG 2012, BGBl. I Nr. 114/2012). Animal housing, treatment, testing and euthanasia were approved by the Styrian government (Amt der Steiermärkischen Landesregierung, Abteilung 13 – Umwelt und Raumordnung Austria).

A total of 16 5xFAD and 16 age matched wild-type (WT) littermates were used for this study, half of the animals were sacrificed for tissue collection at 3 months of age, the other half at 7 months of age. Four males and 4 females were included per group of 8 animals.

5xFAD (Familiar Alzheimer Disease) mice bear five mutations, three in the amyloid precursor protein (APP695) gene [APP K670N/M671L (Swedish), I716V (Florida), V717I (London)] as well as two mutations in the presenilin 1 gene [PS1 M146L, L286V] (25). The expression of the 5xFAD transgene is driven by the neuron specific Thy1 promoter. The five mutations cause an early onset of the cognitive decline and increasing Abeta 1-40 and 1-42 levels in the brain and cerebrospinal fluids, over age. Histological analysis confirmed amyloid plaque load accompanied by neuroinflammation. Thus, the 5xFAD mouse mimics important phenotypic features of amyloidogenic neurodegeneration, neuroinflammation as well as learning and memory deficits.

For sample collection, mice were euthanized by IP injection of 600 mg/kg pentobarbital, dose 1g – 10µl. The thorax was opened and blood was collected by heart puncture with a 23-gauge needle. The needle was removed and the blood was transferred to the sample tube (MiniCollect® K_2_ EDTA (potassium ethylenediaminetetraacetic acid). The tube was inverted thoroughly to facilitate homogeneous distribution of the EDTA and prevent clotting. The blood samples were centrifuged at 3000 x g for 10 minutes at room temperature (22°C). Plasma was transferred to a pre-labeled 1.5ml LoBind Eppendorf tube (1 aliquot 100µl + rest), frozen on dry ice and stored at −80°C.

Animals were transcardially perfused with 0.9% saline. The thoracic aorta - between the lungs and the liver - was clamped with hemostatic forceps to block the blood flow from the heart to the abdomen but allowing the blood flow to the brain. The right atrium was opened with scissors. A constant pressure of 100 to 120 mm Hg was maintained on the perfusion solution by connecting the solution bottle to a manometer-controlled air compressor. Perfusion was continued until the skull surface had turned pale and only perfusion solution instead of blood was exiting of the right atrium.

Thereafter, the skull was opened and afterwards the brain was removed carefully and hemisected on a cooled surface. The left hemibrains were further dissected on a cooled surface into hippocampus, cortex, and rest brain. All parts were weighed, snap frozen on dry ice and stored at −80°C. Cortex and hippocampus were used for downstream evaluations.

### Plasma biomarkers (AD converters - CU and post-mortem BLSA participants)

Blood plasma samples were collected from BLSA participants at the NIA Clinical Research Unit at Harbor Hospital. Details on collection and processing have been described in detail previously (26).

The following 6 plasma biomarkers were assayed: tau phosphorylated at threonine 181 (pTau181), tau phosphorylated at threonine 231 (pTau231), amyloid-β 42 (Aβ-42), Amyloid-β 40 (Aβ-40), glial fibrillary acidic protein (GFAP), and neurofilament light chain (NfL).

Both pTau181 and pTau231 were assayed on an HD-X instrument (Quanterix, Billerica, MA) using Single molecule array (Simoa) assays developed by the clinical laboratory, University of Gothenburg, Mölndal, Sweden. Assay details have been published previously (13, 27). For pTau181, repeatability coefficients and intermediate precision were 9.3% and 11.6% (17.4 pg.mL) and 8.9% and 11.1% (23.8 pg/mL) respectively.

For pTau231, repeatability coefficients and intermediate precision were 8.7% and 13.8% (11.5 pg.mL) and 4.2% and 11.0% (13.7 pg/mL) respectively. Values indicated as below the limit of detection (LOD) were set to 0 (pTau181 n = 1, pTau231 n = 17). Values indicated as below the lower limit of quantification (LLOQ) were used as is (pTau181 n = 10, pTau231 n = 22). Outliers beyond 5x the standard deviation(SD) were set to the 5SD value.

Aβ-42, Aβ-40, GFAP, and NfL were assayed on a Quanterix HD-X instrument using the Quanterix Simoa Neurology 4-plex-E assay. Inter-assay coefficient of variation were as follows: Aβ-40 – 5.2%; Aβ-42 – 5.9%; GFAP – 8.1%; NfL – 7.8%. No values were indicated as either LOD or LLOQ. Outliers beyond 5x the SD were set to the 5SD value. We selected to use the ratio of Aβ-42/Aβ-40 (Aβ ratio) vs. individual measures based on prior work suggesting that a lower plasma ratio is associated with increased risk of developing Alzheimer’s disease (AD) (28, 29).

### Outcomes: MCI/dementia diagnosis

In BLSA, cognitive status is determined at consensus diagnosis conferences; these procedures have been described previously (20). Briefly, consensus conferences include neurologists, neuropsychologists, and neuroimaging scientists who review clinical and neuropsychological data if a participant made four or more errors on the Blessed Information, Memory, and Concentration (BIMC) test, if their Clinical Dementia Rating (CDR) score was equal to or greater than 0.5, or if concerns were raised regarding the patient’s cognitive status by a reliable informant. The diagnosis of dementia and AD are based on the Diagnostic and Statistical Manual (DSM)-III-R and the National Institute of Neurological and Communication Disorders and Stroke-Alzheimer’s Disease and Related Disorders Association (NINCDS-ADRDA) criteria respectively (30). The diagnosis of mild cognitive impairment is based on Petersen criteria (31). For individuals diagnosed with AD or MCI, age at onset of initial symptoms was estimated at consensus case conferences using longitudinal cognitive performance data as well as informant-based history.

### GFAP Plasma, NfL Plasma and GFAP brain assays in 5XFAD transgenic and wild-type mice

Terminal plasma of 5xFAD animals and wildtype littermates was used for analysis of NfL and GFAP levels. For the measurement of NfL and GFAP concentrations in plasma, commercially available ELISA kits were used (NF-light® ELISA 10-7001 CE from UmanDiagnostics; Mouse GFAP Elisa Kit (ab233621) from Abcam). Plasma samples were diluted in kit provided diluents and the assay was performed according to the manual. Plates were read on Cytation 5 multimode reader (Bio-Tek) and values are expressed as pg/mL plasma.

Cortex and hippocampus from 8 animals per group (4 male, 4 female) were homogenized by adding 9 (cortex) or 19 (hippocampus) volumes of tissue homogenization buffer (THB; 250 mM Sucrose, 1% Triton X-100, 1 mM EDTA, 1 mM EGTA, 20 mM Tris pH 7.4) including 1x protease inhibitor (Calbiochem), to cortex or hippocampus samples respectively. The tissue was homogenized with a beadmill (UPHO, Geneye) at 55Hz for 50sec. Both cortex and hippocampus samples were analyzed using R-plex GFAP assay (Mesoscale Discovery, K1511MR) according to the manufacturers protocol. Plates were read on MESO QuickPlex SQ 120MM (Mesoscale Discovery) and values are expressed as pg per mg tissue.

### Outcomes: AD pathology

Postmortem brain examinations were performed by experienced neuropathologists at the Johns Hopkins University School of Medicine; details have been published previously (21). Assessment of neuritic plaques used the CERAD (32) criteria, and assessment of neurofibrillary tangles used Braak (33) criteria.

### Statistical Analysis: AD converters-CU/post-mortem

For the AD converters-CU sample, the primary analytic goal was to test for differences across the 6 plasma biomarkers (5 total measures including Aβ ratio) between AD/MCI (AD converters) and cognitively unimpaired (CU) participants. AD converters had clinical symptoms at onset and were unimpaired at 5 and 10 years prior to onset.

We used separate repeated measure ANCOVA models with each biomarker measure as the outcome. The main predictors for the models included diagnosis group (AD converters vs CU), time bins (10, 5, 0 yrs prior to onset) and diagnosis group x time bins interaction. Covariates included age, age squared, sex, race, and estimated globular filtration rate (eGFR). Various residual covariance structures (that included variance components, compound symmetry, first order autoregressive and unstructured) were tested to determine the best model fit using model fit statistics. This model allowed us to compare the covariates-adjusted means at 10 year prior, 5 year prior and onset by diagnosis group while accounting for within-subject correlations from repeated measurement. We also computed Cohen’s D effect size based on the estimated differences at the three time points.

In order to compare prediction across biomarkers, we used separate logistic regressions to estimate the odds ratio (OR) in predicting AD converters for each biomarker at each time point after adjusting for covariates age, sex, race and eGFR. Biomarkers were median centered and scaled by the interquartile range (IQR) to quantify the odds ratios for each IQR change in the biomarker. The sign of the Aβ ratio was reversed so that its odds ratios were in the same direction as other biomarkers.

In the AD converters-CU sample, we additionally calculated the association between biomarkers (at onset, combining case and control) using partial spearman correlations and controlling for age and sex.

For the AD converters-CU sample, the secondary analytic goal was to test whether concentrations of the plasma biomarkers differed by *APOE* genotype within the AD converters and CU separately. Using the same repeated measure ANCOVA model described above, we added *APOE ε*4 carrier status (excluding *ε*2/*ε*4 subjects); its two way interaction with diagnosis group and time bins; and the three way interaction of *APOE ε*4 x diagnosis group x time bins.

For the postmortem sample, the primary analytic goal was to test whether the 6 plasma biomarkers (5 total measures including Aβ ratio) collected prior to death were associated with postmortem-confirmed clinical diagnosis (AD vs ASY; AD vs CN; ASY vs CN) and severity of AD neuropathology (i.e. CERAD and Braak scores). For analyses of pathology, participants were grouped in 3 groups based on increasing severity: (0,1), 2, 3 for CERAD and (1,2), (3,4), (5,6) for Braak scores.

We used proportional odds ordinal logistic models (biomarker as the outcome), a generalization of the Wilcoxon and Kruskal-Wallis test that allows for covariates, including age, sex and time between the biomarker sample and death. We first tested the global null hypothesis that biomarker levels across all three groups were equal. For biomarkers with significant p-values (p < 0.05) rejecting the global null hypothesis, we then tested the pairwise group comparisons. We additionally evaluated the correlation between CERAD and Braak scores with each biomarker using spearman partial correlations adjusted for age, sex, and the time between the biomarker sample and death.

The analyses were conducted in SAS 9.4 (Cary, NC) and visualized in R (version 4.2.3).

### Statistical Analysis: 5XFAD transgenic and wild-type mice

In the animal studies, the primary analytic goal was to test for differences in brain tissue and plasma GFAP levels between transgenic and wild-type mice, determine correlations between brain GFAP levels and blood plasma GFAP levels, and determine correlations between blood plasma GFAP and blood plasma NfL levels. Brain GFAP and plasma NfL were right skewed and therefore natural log transformed. T-tests were used to test for differences in plasma and brain (cortex and hippocampus) GFAP levels between transgenic and wild-type mice at 3 and 7 months. Pearson correlations were used to determine associations between GFAP brain and GFAP plasma levels separately in transgenic and wild-type mice at 3 and 7 months.

The analyses were conducted in Stata (version 16.0; StataCorp) and visualized in R (version 4.2.1).

## Results

**Table 1.**

| **Table1** | Whole sample | CU | AD Converters (47 MCI and 111 AD) | P values |
| --- | --- | --- | --- | --- |
| N subjects | 318 | 160 | 158 |  |
| Female | 156 (49.1%) | 79 (49.4%) | 77 (48.7%) | 0.91 |
| White | 256 (80.5%) | 127(79.4%) | 129 (81.7%) | 0.61 |
| Black | 54 (17.0%) | 28 (17.5%) | 26 (16.5%) | 0.80 |
| Other race | 8 (2.5%) | 5 (3.1%) | 3 (1.9%) | 0.72 |
| APOE e4 +<br>Note: 5 missing in both) | 82 (26.2%) | 37 (23.4%) | 45 (29.0%) | 0.26 |
| Age<br>10 yr prior to onset | 75.1 (6.7)<br>51.3 – 87.9 | 75.1 (6.7)<br>51.6 – 87.5 | 75.1 (6.7)<br>51.3 – 87.9 | 0.97 |
| Age<br>5 year prior to onset | 80.4 (6.0)<br>58.3 – 93.7 | 80.5 (6.0)<br>58.5 – 92.5 | 80.4 (6.0)<br>58.3 – 93.7 | 0.86 |
| Age<br>at onset or follow-up | 84.8 (5.9)<br>65.8 – 97.4 | 84.5 (5.9)<br>65.8 – 95.7 | 85.2 (6.0)<br>66.0 – 97.4 | 0.36 |
| Total N visits | 756 | 379 | 377 |  |
| N visits 10 years prior | 234 | 117 | 117 |  |
| N visits 5 years prior | 281 | 140 | 141 |  |
| N visits around onset | 241 | 122 | 119 |  |
| N subjects contributing to total visits: |  |  |  |  |
| 1 | 42 (13.2%) | 22 (13.8%) | 20 (12.7%) |  |
| 2 | 114 (35.9%) | 57 (35.6%) | 57 (36.1%) |  |
| 3 | 162 (50.9%) | 81(50.6%) | 81 (51.3%) |  |

Table 1 summarizes demographic characteristics of the AD converters – CU sample (n = 158; n = 160 respectively). Participants were 49.1% female, 80.5% white, and 26.2% were *APOE ε*4 carriers. Due to *a priori* matching/sampling criteria, AD converters and CU did not vary significantly across demographic characteristics. AD converters were aged 85.2 years at MCI/AD symptom onset, and 80.4 and 75.1 years 5 years and 10 years prior respectively. CU were age-matched to AD converters. Approximately 50.9% of participants provided data at all three time points (i.e.,10 years prior, 5 years prior, and at onset of MCI/AD symptoms), 35.9% provided data at two timepoints and 13.2% provided data at only one timepoint.

Figure 1 shows covariate adjusted means for AD converters and CU at each time point across all 5 biomarker measures. Plasma concentration of GFAP was significantly higher in AD converters compared with CU across all three time points (estimated difference (standard error), p-value) at 10 years prior to symptom onset (35.88 pg/mL (9.04), p < 0.0001), 5 years prior to symptom onset (31.53 pg/mL (9.33), p = 0.0008) and at symptom onset (46.09 pg/mL (11.01), p < 0.0001). Aβ ratio was not significant at any of the three timepoints. pTau181, pTau231, and NfL were significantly higher in AD converters compared with CU at symptom onset only (1.63 pg/mL (0.73), p = 0.025); (1.65 pg/mL (1.21), p = 0.049); (3.56 pg/mL (1.65), p = 0.032) respectively. Supplementary Table 1 includes covariate adjusted means for AD converters and CU across all biomarkers at each timepoint.

**Figure 1.**
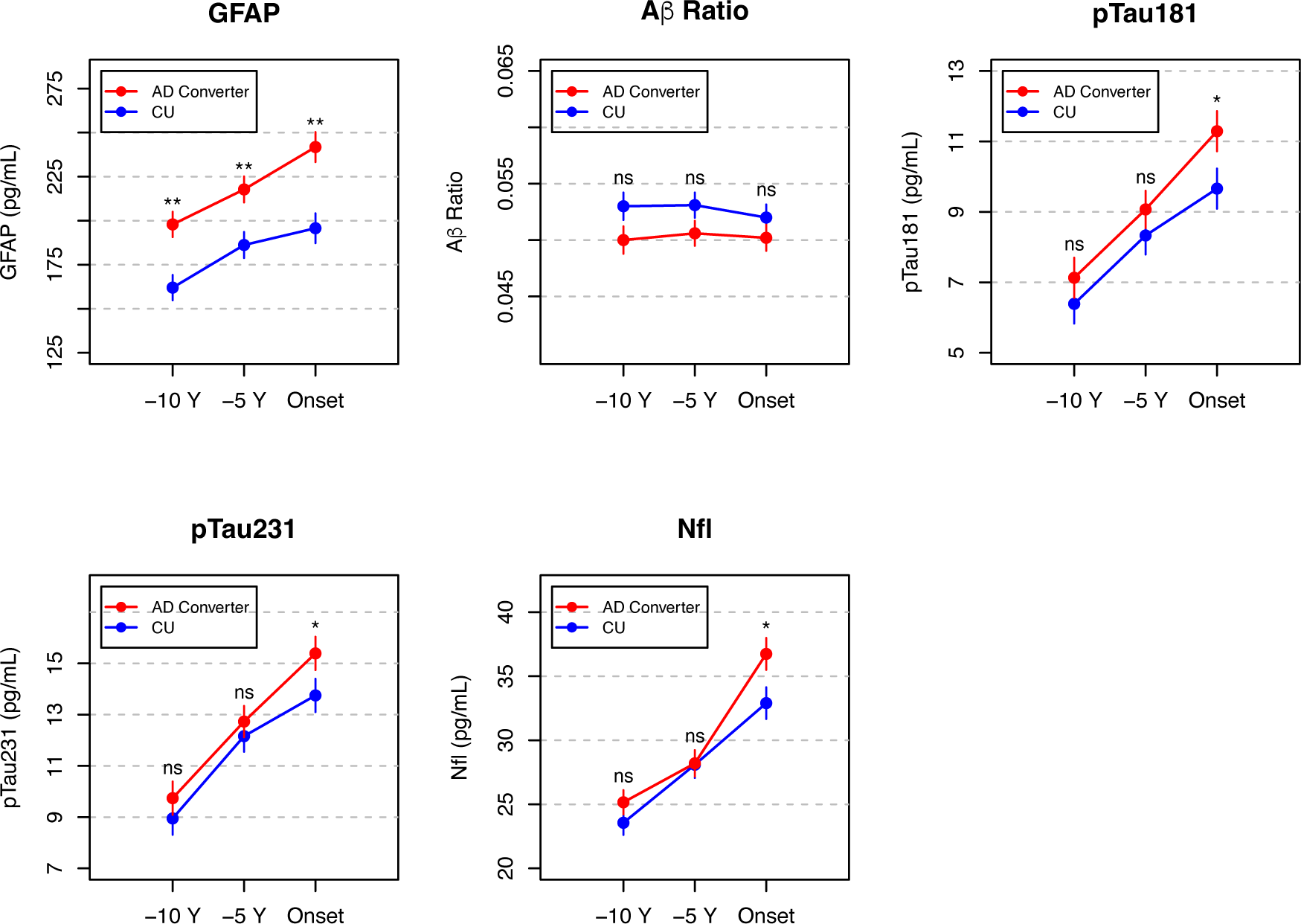
Plasma biomarker differences – AD converters vs Cognitively Unimpaired. Differences across five plasma biomarkers comparing participants who converted to Alzheimer’s disease or MCi (AD converters; n = 158) to participants who remained cognitively unimpaired (CU; n = 160). GFAP was the only plasma biomarker with significant differences between AD converters and CU across all three timepoints. Aβ ratio was not significant at any timepoint; pTau231, pTau181, and NfL were significant at symptom onset only. Serial plasma samples were taken at symptom onset and 5 years (−5 Y) and 10 years (−10 Y) prior to onset. CU were sex, race, age and visit year-matched to AD converters at all three timepoints. Separate repeated measure ANCOVA models were used to compare covariate adjusted means at all timepoints. NS: not significant (p > 0.05); * p < 0.05; ** p < 0.01; Alzheimer’s disease (AD); Cognitively Unimpaired (CU); Glial Fibrillary Acidic Protein (GFAP); Neurofilament-Light chain (NfL); Beta-Amyloid (Aβ); Aβ-42/Aβ-40 (Aβ ratio); Phosphorylated Tau (p-tau)

Figure 2 summarizes the odds ratio (OR) and 95% confidence intervals (CI) across all biomarkers at all 3 time points; GFAP was the only biomarker to significantly predict AD converters vs CU at all three time points including 10 years prior to symptom onset (OR = 2.24, p = 0.0013); 5 years prior to symptom onset (OR = 1.56, p = 0.0062); and at onset (OR = 1.50, p < 0.0001). Supplementary Table 2 incudes covariate adjusted OR and 95% confidence intervals (CI) across all biomarkers at each time point.

**Figure 2.**
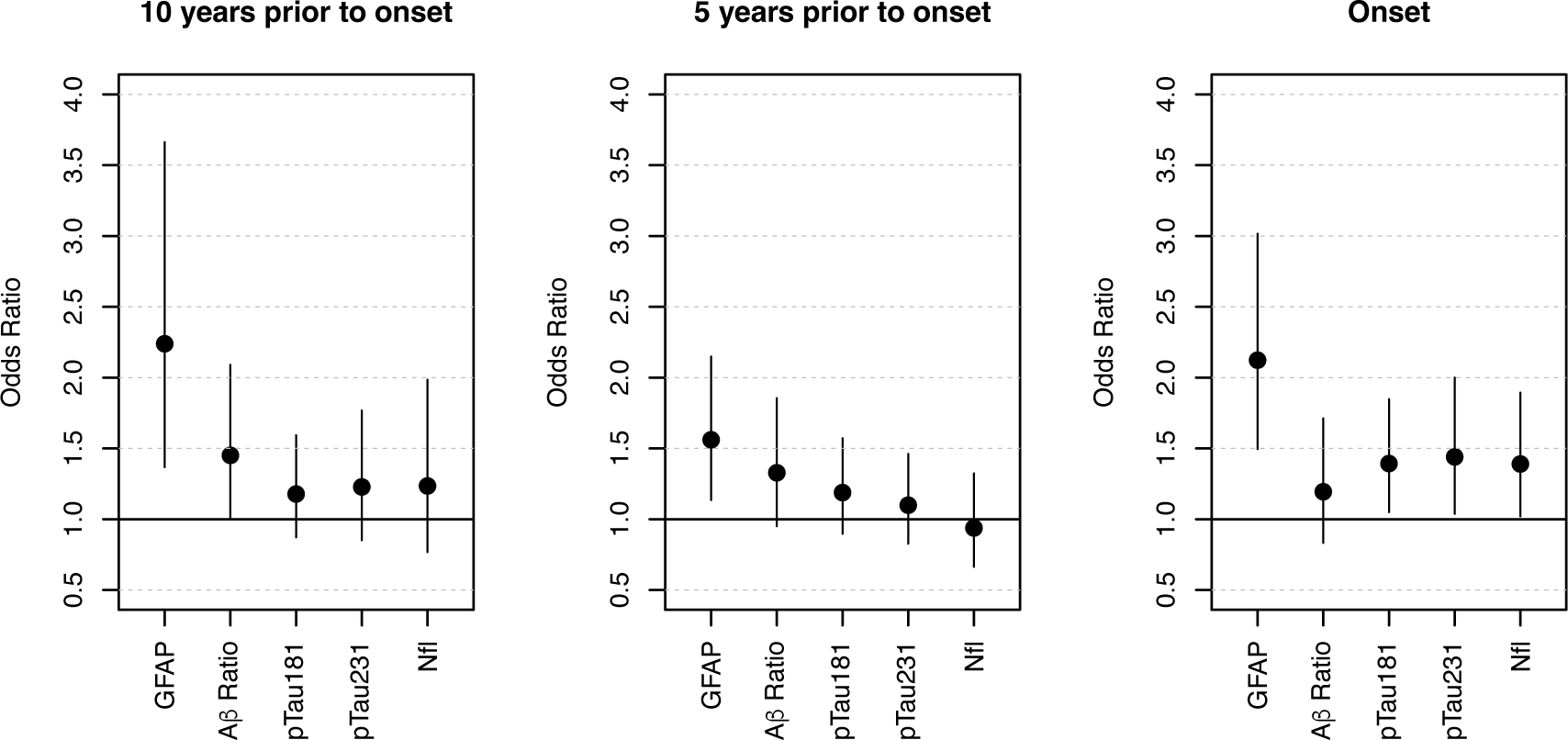
Plasma biomarker prediction of AD converters vs CU. Odds Ratio (OR) and 95% confidence intervals across five plasma biomarkers for predicting AD converters vs CU. GFAP was the only plasma biomarker to significantly predict AD converters vs CU at all three time points. Separate logistic regression were used to estimate OR. The sign of the Aβ ratio was reversed so that its odds ratios were in the same direction as other biomarkers for ease of visualization/ interpretation. Alzheimer’s disease (AD); Cognitively Unimpaired (CU); Glial Fibrillary Acidic Protein (GFAP); neurofilament-light chain (NfL); Beta-Amyloid (Aβ); Aβ-42/Aβ-40 (Aβ ratio); Phosphorylated Tau (p-tau)

Partial spearman correlations among all plasma biomarkers at symptom onset showed that the highest correlation with GFAP was for NfL (r = 0.50), followed by correlations with pTau181 (r = 0.34) and pTau231 (r = 0.35). The highest correlation with Aβ ratio was with GFAP (r = −0.25). For NfL, the highest correlation was also with GFAP; pTau181 and pTau231 had the highest correlations across all comparisons (r = 0.74). Supplementary Table 3 includes all pairwise partial spearman correlations.

In secondary analyses to test whether plasma concentrations of the AD biomarkers differed by *APOE* genotype within the AD converter and CU groups (Supplementary Table 4), we observed that plasma GFAP levels were significantly higher in *APOE ε4* carriers compared to non-carriers among AD converters 5 years prior to symptom onset ( P= 0.0149) with a similar trend at the onset of symptoms (P = 0.0731). There were no differences in GFAP levels comparing *APOE ε4* carriers to non-carriers among CU. Plasma pTau231 was higher in *APOE ε*4 carriers compared to non-carriers among AD converters at all three time points (10 year prior: p = 0.0044, 5 years prior: p = .0043, onset p = 0.0009. These trends were not observed among CU.

Table 2 describes demographic characteristics of the postmortem study (n = 65). Age at death was 82.6 (CN), 87.0 (ASY), and 88.1 years (AD). The approximate number of years (i.e., interval) between blood draw and death was 3.5 (CN), 2.9 (ASY), 5.8 years (AD). 44.6% of participants were female, 95.4% white, and 27.7% were *APOE ε*4 carriers. The three participant groups (CN, ASY, AD) significantly varied in the interval between age at blood draw and death, and percent White.

**Table 2.**

| **Table 2** | Whole sample | CN | ASY | AD | P value* |
| --- | --- | --- | --- | --- | --- |
| N | 65 | 19 | 18 | 28 |  |
| Age death | 86.2 (10.0)<br>52.9 – 103.1 | 82.6 (13.0)<br>52.9 – 102.2 | 87.0 (7.9)<br>71.9 – 96.4 | 88.1 (8.4)<br>62.9 – 103.1 | 0.42 |
| Age at blood draw | 81.9 (11.1)<br>44.1 – 99.1 | 79.1 (14.3)<br>44.1 – 97.2 | 84.1 (8.3)<br>64.2 – 92.4 | 82.3 (10.1)<br>52.9 – 99.1 | 0.66 |
| Interval | 4.3 (3.4)<br>0.2 – 15.6 | 3.5 (2.6)<br>0.5 – 8.9 | 2.9 (2.4)<br>0.5 – 7.7 | 5.8 (3.9)<br>0.2 – 15.6 | 0.015 |
| Female | 29 (44.6%) | 6 (31.6%) | 8 (44.4%) | 15 (53.6%) | 0.35 |
| White | 62 (95.4%) | 16 (84.2%) | 18 (100%) | 28 (100%) | 0.041 |
| *APOE* $\epsilon 4$ | 18 (27.7%) | 5 (26.3%) | 5 (27.8%) | 8 (28.6%) | 1.00 |

Plasma GFAP and pTau231 were the only two biomarkers with significant differences (global p < 0.05) in concentration across all comparisons i.e.: postmortem-confirmed clinical diagnosis and severity of AD neuropathology, including CERAD and Braak scores. Considering pairwise differences, GFAP was the only biomarker that showed significant differences between all clinical diagnosis comparisons (AD vs CN; ASY vs CN; and AD vs ASY) (Figure 3). pTau181 and pTau231 showed significant differences between AD vs CN and AD vs ASY and Aβ ratio showed a significant difference between AD vs CN.

**Figure 3.**
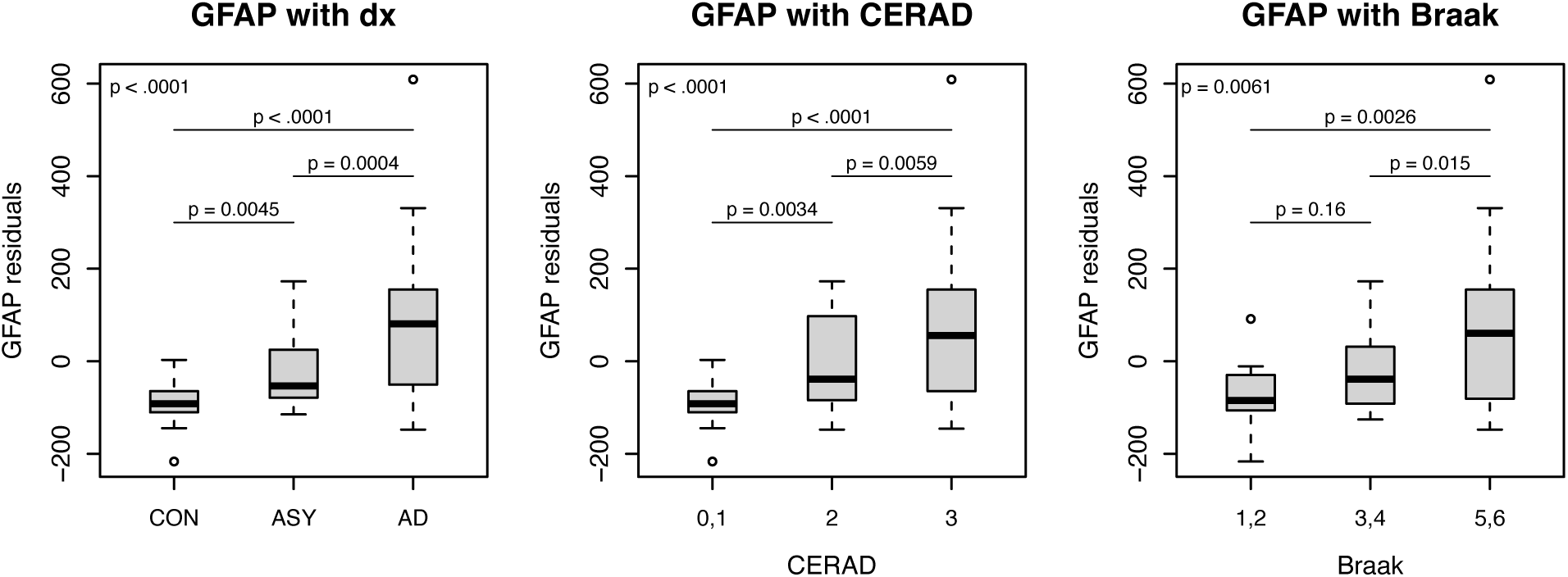
Plasma GFAP differences by clinical diagnosis and severity of AD pathology. Plasma GFAP differences across three comparisons: postmortem-confirmed clinical diagnosis (dx) and severity of AD neuropathology assessed by CERAD and Braak scores. Global p-values (top left corner) and pair-wise p-values generated using proportional odds ordinal logistic models. Diagnosis (dx); Glial Fibrillary Acidic Protein (GFAP); Consortium to Establish a Registry for Alzheimer’s disease (CERAD); Control (CON); asymptomatic Alzheimer’s disease (ASY); Alzheimer’s Disease (AD)

GFAP was the only biomarker that showed significant differences between all CERAD score comparisons (3 vs 0,1; 2 vs 0,1; 3 vs 2) (Figure 3). pTau231 showed significant differences between 3 vs 0,1 and 3 vs 2, pTau181 showed a significant difference between 3 vs 0,1, and Aβ ratio showed a significant differences between 3 vs 0,1 and 2 vs 0,1.

GFAP, as well as pTau181 and pTau231, had similar difference patterns across Braak score groups (p < 0.05 for 5,6 vs 1,2; 5,6 vs 3,4) with higher biomarker levels associated with higher Braak scores (Figure 3). Aβ ratio did not show significant differences between Braak score groups.

NfL did not show significant differences across any of the three comparisons (clinical diagnosis, CERAD and Braak scores). Supplementary Table 5 and Supplementary Figure 1 includes global and pairwise p-values across biomarkers.

Results from spearman partial correlation analyses between biomarkers and CERAD and Braak scores were complementary to the results described above and in Supplementary Table 4. GFAP was significantly correlated with both CERAD and Braak scores (r = 0.60 and r= 0.43 respectively). pTau231 and pTau181 had the highest correlation with Braak score (r = 0.58 and r = 0.43 respectively) and were also correlated with CERAD score (r = 0.37 and r = 0.31 respectively). Aβ ratio was significantly correlated only with CERAD score (r = −0.32), and NfL was not significantly correlated with either CERAD or Braak scores. Supplementary Table 6 includes correlations and p-values across biomarkers.

Brain GFAP levels in the hippocampus were significantly higher in 5xFAD transgenic mice compared to WT at 3 months and 7 months, whereas brain GFAP levels in the cortex were significantly higher in transgenic mice compared to wildtype at 7 months (Figure 4A). While there were no statistically significant (p < 0.05) differences in plasma GFAP levels between the 5xFAD transgenic mice compared to WT at 3 months or 7 months of age, plasma GFAP and brain cortex GFAP levels were highly correlated (r = -.77, p = 0.027) in transgenic mice at 7 months (Figure 4B). Supplementary Table 7 includes mean differences and correlations across all comparisons between 5xFAD and WT mice.

**Figure 4.**
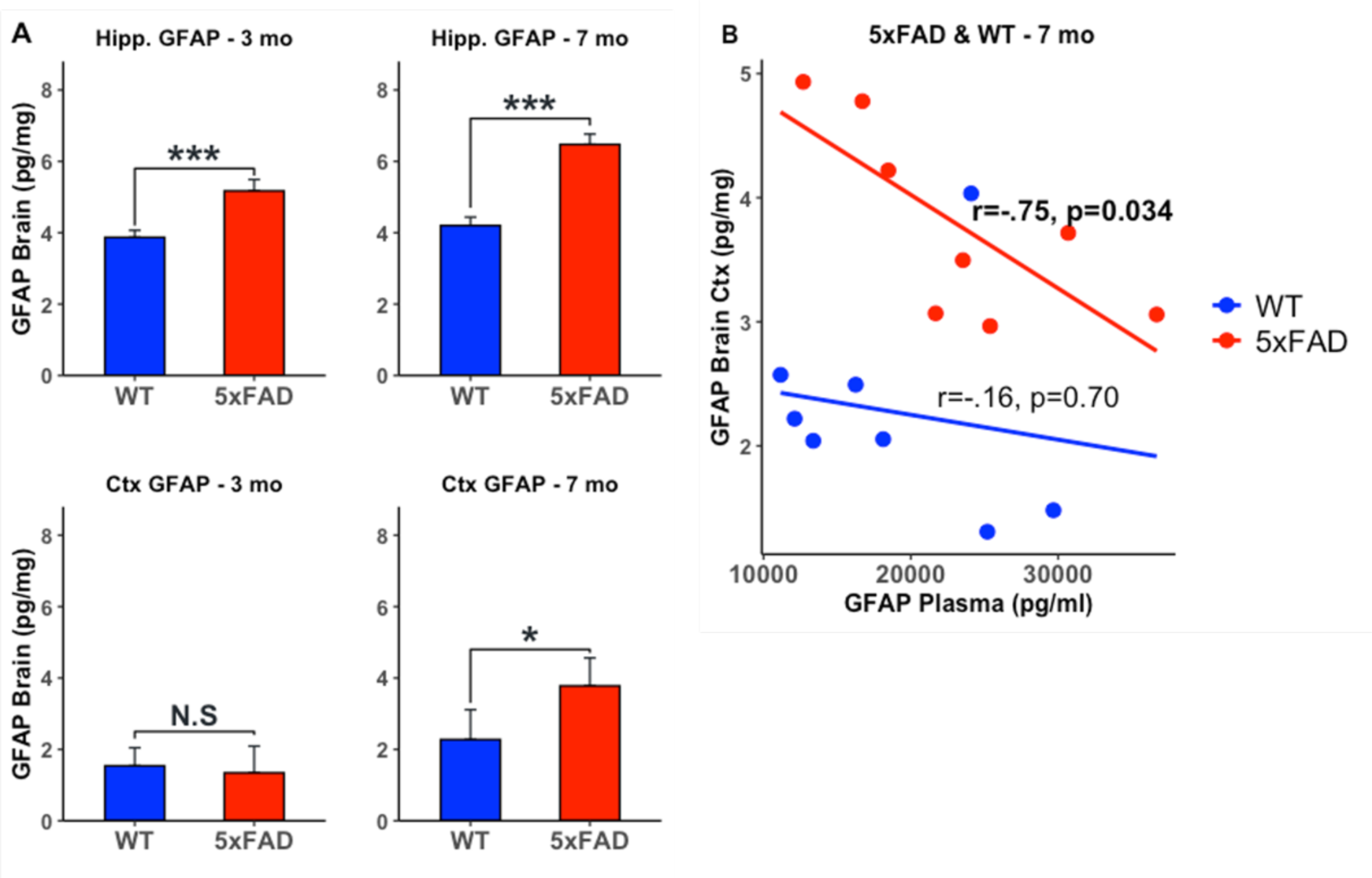
Brain and plasma GFAP levels in WT and 5xFAD mice. A. Brain and plasma GFAP differences in WT and 5xFAD mice. Brain GFAP levels in the hippocampus were significantly higher in 5xFAD mice compared to WT at 3 and 7 mo and significantly higher in the cortex at 7 mo. B. Plasma GFAP and Brain GFAP levels were significantly negatively correlated at 7 mo in 5xFAD mice. Wild Type (WT); 5x Familial Alzheimer’s Disease (5xFAD); Glial Fibrillary Acidic Protein (GFAP); Cortex (Ctx); Hippocampus (Hipp); Months (mo); NS: not significant (p > 0.05); * p < 0.05; *** p < 0.001;

## Discussion

The main findings of this study are that increased astrocyte reactivity (34), assessed by elevated plasma GFAP levels is an early event in the temporal progression of blood biomarker changes in preclinical AD. In AD-converters who are initially cognitively unimpaired, we find that higher plasma GFAP levels are observed as early as 10-years prior to the onset of cognitive impairment due to incident AD compared to those who remain cognitively unimpaired (i.e. in CU). Plasma GFAP levels in AD-converters remain elevated 5-years prior to as well as coincident with the onset of cognitive impairment due to incident AD.

While reductions in plasma Aβ-42/Aβ-40 (Aβ ratio) were not observed across all timepoints, biomarkers that are believed to reflect neurodegeneration - i.e., pTau231, pTau181 and NfL (35) - are increased in AD-converters only at the onset of cognitive impairment relative to CU individuals. Furthermore, in participants with neuropathologically confirmed AD at autopsy, we observed that plasma GFAP levels are elevated relative to cognitively normal (CN) individuals and intermediate in those who remain cognitively unimpaired despite significant levels of AD pathology (ASYMAD). We also confirmed that higher plasma GFAP levels at death are associated with greater severity of both pathological hallmarks of AD, i.e., neuritic plaques and neurofibrillary tangles.

Together, these results implicate astrocyte reactivity as a key upstream perturbation in the cascade of AD pathogenesis that may be detectable before changes in proxies of accumulation of amyloid pathology and neurodegeneration in individuals at risk for AD. Our findings are consistent with a recent study from human post-mortem samples that reported a higher density of GFAP-positive astrocytes in AD compared to control brains (e.g., (36)). While it remains to be established whether astrocyte reactivity is an early causal driver of AD, the associations of elevated plasma GFAP levels with severity of both neuritic plaque and neurofibrillary pathology as well as with pTau and NfL levels suggest that it may be important both as a potential initiator of AD pathogenesis as well as a plausible mediator of intermediate and late stages of AD progression. This hypothesis is also supported by recent findings by Bellaver and colleagues who showed that astrocyte reactivity measured by elevated GFAP levels is an important upstream event linking Aβ deposition in the brain with accumulation of tau pathology in individuals at risk for AD (37). Our findings that plasma GFAP levels in ASYMAD participants are intermediate between AD and CN groups further suggest that astrocyte reactivity may be an important determinant of symptom onset in individuals with AD pathology. In this context, it is especially interesting to note that we recently showed increased levels of STAT3 protein, a key regulator of GFAP expression (38, 39) in the brains of young *APOE* ε4 carriers, decades before the typical age at onset of sporadic AD (40), suggesting that increased astrocyte reactivity may be an early driver of AD pathogenesis. This hypothesis is further supported by our current results showing that cognitively normal *APOE ε*4 carriers who subsequently progress to AD have higher levels of plasma GFAP compared to *APOE ε*4 non-carriers.

The increase in plasma GFAP levels in AD is thought to reflect its release from astrocyte end feet into capillaries in the brain (41). However, few studies have examined whether there is a direct relationship between brain and plasma GFAP concentrations. We addressed this question in the 5xFAD transgenic AD model and observed a significant increase in both cortical and hippocampal GFAP concentrations in transgenic mice at 7 months of age compared to wild-type mice, a time point preceding the onset of spatial memory impairment that is typically observed at 9 months in this AD model (42). While we did not observe significant differences in plasma GFAP levels between transgenic and wild-type mice at either time point, we observed a strong negative correlation between cortical and plasma GFAP levels in transgenic mice at 7 months of age. These results are similar to a recent study by Chiotis et al. who found a negative correlation between plasma GFAP concentrations and a PET biomarker of reactive astrocytosis; ^11^C-deuterium-L-deprenyl (^11^C-DED), in patients with sporadic AD (43). The authors suggest that the divergence between ^11^C-DED and plasma GFAP may reflect distinct states of reactive astrocytosis measured by these biomarkers. While the precise mechanisms underlying the divergence in plasma and brain GFAP concentrations remain to be established, several additional factors merit consideration including the contribution of peripheral tissues such as liver, pancreas, lymphocytes and cartilage to measured plasma GFAP levels (39, 44).

Key strengths of this study include serial measurements of six AD blood biomarkers in a well characterized longitudinal sample of older individuals with up to a 10-year follow up interval, inclusion of a post-mortem cohort with quantification of AD pathology and confirmation of clinical diagnosis, as well as quantification of plasma and brain GFAP levels in a well-characterized transgenic AD mouse model.

Limitations include the relatively small sample size in the BLSA autopsy cohort and lack of GFAP measures from these brain tissue samples. The accuracy of plasma Aβ immunoassays to detect brain amyloid pathology in our study may also be lower than mass spectrometry-based approaches (45). Our results also suggest that changes in plasma GFAP levels may begin even earlier than 10 years prior to the onset of cognitive impairment in patients who eventually develop AD. This is a hypothesis that remains to be confirmed in plasma samples collected even earlier than those in the present study.

Our findings have important implications. They suggest that reactive astrocytosis, an established biological response to neuronal injury (41, 46) may be an early initiator of AD pathogenesis, preceding brain amyloid accumulation, tau deposition, neurodegeneration and the onset of cognitive impairment in patients who eventually develop AD. Together with the observation that plasma GFAP levels are associated both with severity of AD pathology and manifestation of AD symptoms, they indicate that experimental treatments targeting reactive astrocytosis may be promising candidate AD therapies. We recently showed that the commonly used immune modulator drug hydroxychloroquine, inactivates STAT3, a key regulator of GFAP expression and ameliorates molecular abnormalities relevant to AD (47). Furthermore, we demonstrated that hydroxychloroquine rescues impaired hippocampal synaptic plasticity in the APP/PS1 mouse model of AD as well as reduces risk of incident AD in a large real world clinical dataset (47). Monitoring plasma GFAP levels as a surrogate outcome measure in studies of experimental AD treatments may be a useful strategy to determine their potential utility as disease-modifying AD treatments.

In summary, we demonstrate that plasma levels of GFAP, a measure of reactive astrocytosis in the brain, are increased a decade prior to the onset of cognitive impairment in AD. The associations of plasma GFAP concentrations with severity of AD pathology and onset of AD symptoms suggest that reactive astrocytosis may be an important mediator of disease progression and a promising target of novel AD treatments.

## Data Availability

Baltimore Longitudinal Study of Aging (BLSA) data are available to researchers and can be requested at https://www.blsa.nih.gov/researchers.

## Acknowledgements

This work was supported in part by the Intramural Research Program of the National Institute on Aging. We are grateful to the participants in the Baltimore Longitudinal Study.

JT is supported by the Johns Hopkins Alzheimer’s Disease Center (P30AG066507).

HZ is a Wallenberg Scholar supported by grants from the Swedish Research Council (#2023-00356; #2022-01018 and #2019-02397), the European Union’s Horizon Europe research and innovation programme under grant agreement No 101053962, Swedish State Support for Clinical Research (#ALFGBG-71320), the Alzheimer Drug Discovery Foundation (ADDF), USA (#201809-2016862), the AD Strategic Fund and the Alzheimer’s Association (#ADSF-21-831376-C, #ADSF-21-831381-C, and #ADSF-21-831377-C), the Bluefield Project, the Olav Thon Foundation, the Erling-Persson Family Foundation, Stiftelsen för Gamla Tjänarinnor, Hjärnfonden, Sweden (#FO2022-0270), the European Union’s Horizon 2020 research and innovation programme under the Marie Skłodowska-Curie grant agreement No 860197 (MIRIADE), the European Union Joint Programme – Neurodegenerative Disease Research (JPND2021-00694), the National Institute for Health and Care Research University College London Hospitals Biomedical Research Centre, and the UK Dementia Research Institute at UCL (UKDRI-1003).

## Conflicts of interest

HZ has served at scientific advisory boards and/or as a consultant for Abbvie, Acumen, Alector, Alzinova, ALZPath, Annexon, Apellis, Artery Therapeutics, AZTherapies, Cognito Therapeutics, CogRx, Denali, Eisai, Merry Life, Nervgen, Novo Nordisk, Optoceutics, Passage Bio, Pinteon Therapeutics, Prothena, Red Abbey Labs, reMYND, Roche, Samumed, Siemens Healthineers, Triplet Therapeutics, and Wave, has given lectures in symposia sponsored by Alzecure, Biogen, Cellectricon, Fujirebio, Lilly, and Roche, and is a co-founder of Brain Biomarker Solutions in Gothenburg AB (BBS), which is a part of the GU Ventures Incubator Program (outside submitted work).

## Notes

### Funding Statement

This work was supported in part by the Intramural Research Program of the National Institute on Aging.
JT is supported by the Johns Hopkins Alzheimers Disease Center (P30AG066507).
HZ is a Wallenberg Scholar supported by grants from the Swedish Research Council (#2023-00356; #2022-01018 and #2019-02397), the European Unions Horizon Europe research and innovation programme under grant agreement No 101053962, Swedish State Support for Clinical Research (#ALFGBG-71320), the Alzheimer Drug Discovery Foundation (ADDF), USA (#201809-2016862), the AD Strategic Fund and the Alzheimers Association (#ADSF-21-831376-C, #ADSF-21-831381-C, and #ADSF-21-831377-C), the Bluefield Project, the Olav Thon Foundation, the Erling-Persson Family Foundation, Stiftelsen for Gamla Tjanarinnor, Hjarnfonden, Sweden (#FO2022-0270), the European Unions Horizon 2020 research and innovation programme under the Marie Sklodowska Curie grant agreement No 860197 (MIRIADE), the European Union Joint Programme Neurodegenerative Disease Research (JPND2021-00694), the National Institute for Health and Care Research University College London Hospitals Biomedical Research Centre, and the UK Dementia Research Institute at UCL (UKDRI-1003).

### Author Declarations

The BLSA study protocol has ongoing approval from the Institutional Review Board of, National Institutes of Health

