## Supplemental Tables & Figures for "Longitudinal progression of blood biomarkers reveals a key role of astrocyte reactivity in preclinical Alzheimer’s disease"

**Supplementary Table 1.** Covariates adjusted means (standard errors) for AD converters, CU and their differences across all biomarkers at each timepoint.

| | Time | Control | Case | difference (Case – control) | Effect size |
|---|---|---|---|---|---|
| GFAP | 10 years prior | 162.00 (7.20) | 197.88(7.20) | 35.88 (9.04)<br>P < .0001 | 0.52 |
| | 5 years prior | 186.21 (7.40) | 217.74 (7.38) | 31.53 (9.33)<br>P = 0.0008 | 0.34 |
| | Onset | 195.72 (8.48) | 241.80 (8.52) | 46.09 (11.01)<br>P < .0001 | 0.47 |
| AB ratio | 10 years prior | 0.053 (0.0012) | 0.050 (0.0012) | -0.0030(0.0016)<br>P = 0.062 | -0.25 |
| | 5 years prior | 0.053 (0.0011) | 0.051 (0.0011) | -0.0025 (0.0014)<br>P = 0.074 | -0.23 |
| | Onset | 0.052 (0.0012) | 0.050 (0.0012) | -0.0018 (0.0015)<br>P = 0.21 | -0.16 |
| pTau181 | 10 years prior | 6.39 (0.56) | 7.13 (0.57) | 0.74 (0.73)<br>P = 0.31 | 0.13 |
| | 5 years prior | 8.33 (0.54) | 9.07 (0.53) | 0.75 (0.69)<br>P = 0.28 | 0.13 |
| | Onset | 9.66 (0.57) | 11.29 (0.57) | 1.63 (0.73)<br>P = 0.025 | 0.25 |
| pTau231 | 10 years prior | 8.95 (0.64) | 9.74 (0.65) | 0.80 (0.83)<br>P = 0.34 | 0.14 |
| | 5 years prior | 12.16 (0.61) | 12.73 (0.61) | 0.57 (0.78)<br>P = 0.47 | 0.081 |
| | Onset | 13.75 (0.65) | 15.39 (0.65) | 1.65 (0.83)<br>P = 0.049 | 0.23 |
| Nfl | 10 years prior | 23.56 (0.95) | 25.16 (0.95) | 1.61 (1.21)<br>P = 0.19 | 0.16 |
| | 5 years prior | 28.09 (1.03) | 28.20 (1.03) | 0.11 (1.33)<br>P = 0.93 | 0.0089 |
| | Onset | 32.90 (1.24) | 36.47 (1.25) | 3.56 (1.65)<br>P = 0.032 | 0.22 |

Model covariates included age, sex, race and eGFR
Alzheimer's disease (AD); Cognitively Unimpaired (CU); Glial Fibrillary Acidic Protein (GFAP); Neurofilament-Light chain (NfL); Beta-Amyloid (Aβ); Aβ-42/Aβ-40 (Aβ ratio); Phosphorylated Tau (p-tau)

**Supplementary Table 2.** Covariates adjusted odds ratios (per IQR change in biomarker) in predicting AD converters for each biomarker at each time point:

| OR (95% CI) | 10 year prior | 5 year prior | Onset |
|---|---|---|---|
| GFAP | 2.24 (1.37, 3.66) P = 0.0013 | 1.56 (1.14, 2.15) P = 0.0062 | 2.12 (1.50, 3.02) P < 0.0001 |
| Ab ratio* | 1.45 (1.01, 2.09) P = 0.0453 | 1.33 (0.951, 1.86) P = 0.0953 | 1.19 (0.833, 1.71) P = 0.335 |
| Ptau181 | 1.18 (0.872, 1.59) P = 0.286 | 1.19 (0.897, 1.57) P = 0.229 | 1.39 (1.05, 1.85) P = 0.0215 |
| Ptau231 | 1.23 (0.852, 1.77) P = 0.270 | 1.10 (0.827, 1.46) P = 0.516 | 1.44 (1.04, 2.00) P = 0.0291 |
| Nfl | 1.24 (0.768, 1.99) P = 0.385 | 0.938 (0.664, 1.32) P = 0.716 | 1.39 (1.02, 1.89) P = 0.0367 |

*The sign of the Aβ ratio was reversed so that its odds ratios were in the same direction as other biomarkers for ease of visualization/ interpretation.
Model covariates included age, sex, race and eGFR.

Alzheimer's disease (AD); Cognitively Unimpaired (CU); Glial Fibrillary Acidic Protein (GFAP); neurofilament-light chain (NfL); Beta-Amyloid (Aβ); Aβ-42/Aβ-40 (Aβ ratio); Phosphorylated Tau (p-tau); Odds ratio (OR); Confidence Interval (CI)

**Supplementary Table 3.** Partial spearman correlations among all plasma biomarkers at AD symptom onset adjusting for age and sex

| Rho<br>P value | GFAP | AB ratio | Nfl | pTau181 | pTau231 |
|---|---|---|---|---|---|
| GFAP | 1 | -0.25<br><.0001 | 0.50<br><0.0001 | 0.34<br><0.0001 | 0.35<br><0.0001 |
| AB ratio | | 1 | -0.10<br>0.12 | -0.16<br>0.019 | -0.13<br>0.048 |
| Nfl | | | 1 | 0.36<br><0.0001 | 0.43<br><.0001 |
| pTau181 | | | | 1 | 0.74<br>< .0001 |
| pTau231 | | | | | 1 |

Model covariates included age and sex.

Alzheimer's disease (AD); Cognitively Unimpaired (CU); Glial Fibrillary Acidic Protein (GFAP); neurofilament-light chain (NfL); Beta-Amyloid (Aβ); Aβ-42/Aβ-40 (Aβ ratio); Phosphorylated Tau (p-tau)

**Supplementary Table 4.** Covariates adjusted means and differences in plasma biomarkers by APOE genotype within AD converter and CU groups

| | | CU | | | AD converter | | |
|---|---|---|---|---|---|---|---|
| | | *APOE e4-* | *APOE e4+* | difference | *APOE e4-* | *APOE e4+* | difference |
| GFAP | -10 years | 163.40 (8.23) | 158.88 (14.39) | -4.51 (15.76) P = 0.775 | 190.38 (8.37) | 213.77 (13.32) | 23.40 (15.05) P = 0.121 |
| | -5 years | 184.93 (8.21) | 180.79 (15.50) | -4.13 (16.79) P = 0.806 | 207.14 (8.53) | 244.37 (13.39) | 37.24 (15.23) P= 0.0149 |
| | onset | 194.71 (9.49) | 204.69 (18.69) | 9.98 (20.34) P = 0.624 | 233.33 (9.94) | 266.51 (16.24) | 33.19 (18.46) P = 0.0731 |
| Ab ratio | -10 years | 0.0532 (0.00136) | 0.0507 (0.00251) | -0.00254 (0.00279) P = 0.363 | 0.0513 (0.0014) | 0.0468 (0.00221) | -0.00447 (0.00251) P = 0.075 |
| | -5 years | 0.0539 (0.00124) | 0.0508 (0.00221) | -0.00306 (0.00245) P = 0.211 | 0.0514 (0.00126) | 0.0491 (0.00194) | -0.00234 (0.00222) P = 0.293 |
| | onset | 0.0532 (0.00127) | 0.0471 (0.00243) | -0.00607 (0.00266) P = 0.0228 | 0.0508 (0.0013) | 0.0496 (0.00212) | -0.00117 (0.00238) P = 0.625 |
| pTau181 | -10 years | 6.47 (0.62) | 5.58 (1.07) | -0.88 (1.19) P = 0.458 | 6.95 (0.63) | 7.50 (1.03) | 0.55 (1.17) P = 0.635 |
| | -5 years | 8.42 (0.59) | 7.98 (1.06) | -0.44 (1.17) P = 0.707 | 8.65 (0.60) | 10.25 (0.97) | 1.60 (1.09) P = 0.144 |
| | onset | 9.72 (0.61) | 9.24 (1.16) | -0.48 (1.27) P=0.703 | 10.66 (0.64) | 12.92 (1.04) | 2.26 (1.18) P = 0.0556 |
| pTau231 | -10 years | 9.32 (0.72) | 8.27 (1.23) | -1.05 (1.37) P = 0.446 | 8.98 (0.73) | 11.73 (1.20) | 2.75 (1.36) P = 0.044 |
| | 5 years | 12.27 (0.68) | 11.69 (1.22) | -0.58 (1.35) P = 0.669 | 11.67 (0.69) | 15.30 (1.11) | 3.63 (1.26) P = 0.0043 |
| | onset | 13.80 (0.70) | 13.47 (1.35) | -0.33 (1.48) P = 0.823 | 14.23 (0.74) | 18.82 (1.21) | 4.59 (1.37) P = 0.0009 |
| Nfl | -10 years | 23.03 (1.10) | 24.42 (1.87) | 1.39 (2.07) P = 0.505 | 24.74 (1.10) | 25.29 (1.78) | 0.54 (2.01) P = 0.787 |
| | -5 years | 28.32 (1.14) | 24.45 (2.24) | -3.87 (2.43) P = 0.113 | 27.44 (1.19) | 28.98 (1.88) | 1.55 (2.15) P=0.473 |
| | onset | 33.09 (1.41) | 31.93 (2.83) | -1.16 (3.09) P = 0.709 | 35.97 (1.48) | 37.02 (2.45) | 1.05 (2.80) P = 0.707 |

Model covariates included age, sex, race and eGFR.

Alzheimer's disease (AD); Cognitively Unimpaired (CU); Glial Fibrillary Acidic Protein (GFAP); neurofilament-light chain (NfL); Beta-Amyloid (Aβ); Aβ-42/Aβ-40 (Aβ ratio); Phosphorylated Tau (p-tau); APOE ε4- (e4-); APOE ε4+ (e4+)

**Supplementary Table 5.** Biomarker associations with postmortem-confirmed clinical diagnosis and severity of AD neuropathology

Table: biomarkers differences among dx group after adjusting for covariates

| | Global Chi-square (DF) | Global p value | AD vs CN P value | ASY vs CN P value | AD vs ASY P value |
|---|---|---|---|---|---|
| Ab ratio | 8.38 (2) | 0.015 | 0.0038 | 0.106 | 0.178 |
| GFAP | 34.36 (2) | <.0001 | <.0001 | 0.0045 | 0.0004 |
| Nfl | 4.63 (2) | 0.099 | * | * | * |
| pTau181 | 10.92 (2) | 0.0043 | 0.0021 | 0.56 | 0.012 |
| pTau231 | 11.19 (2) | 0.0037 | 0.0036 | 0.94 | 0.0045 |

Table: biomarkers differences among Braak group after adjusting for covariates

| | Global Chi-square (DF) | Global p value | Braak 5,6 vs 1,2 P value | Braak 3,4 vs 1,2 P value | Braak 5,6 vs 3,4 P value |
|---|---|---|---|---|---|
| Ab ratio | 3.22 (2) | 0.20 | * | * | * |
| GFAP | 10.19 (2) | 0.0061 | 0.0026 | 0.16 | 0.015 |
| Nfl | 0.71 (2) | 0.70 | * | * | * |
| pTau181 | 14.07 (2) | 0.0009 | 0.015 | 0.71 | 0.0003 |
| pTau231 | 28.77 (2) | <.0001 | <.0001 | 0.58 | <.0001 |

Table: biomarkers differences among CERAD group after adjusting for covariates

| | Global Chi-square (DF) | Global p value | CERAD 3 vs 0,1 P value | CERAD 2 vs 0,1 P value | CERAD 3 vs 2 P value |
|---|---|---|---|---|---|
| Ab ratio | 6.71 (2) | 0.035 | 0.015 | 0.041 | 0.70 |
| GFAP | 29.19 (2) | <.0001 | <.0001 | 0.0034 | 0.0059 |
| Nfl | 0.37 (2) | 0.83 | * | * | * |
| pTau181 | 6.52 (2) | 0.038 | 0.011 | 0.25 | 0.16 |
| pTau231 | 8.38 (2) | 0.015 | 0.0059 | 0.70 | 0.022 |

* pair-wise differences were not calculated where the global p-value was not significant (p > 0.05).

Alzheimer's disease (AD); Cognitively Unimpaired (CU); Glial Fibrillary Acidic Protein (GFAP); neurofilament-light chain (NfL); Beta-Amyloid (Aβ); Aβ-42/Aβ-40 (Aβ ratio); Phosphorylated Tau (p-tau); diagnosis (dx); Consortium to Establish a Registry for Alzheimer's disease (CERAD); degrees of freedom (DF); diagnosis (dx)

**Supplementary Figure 1.** Box plots of plasma biomarker differences by postmortem-confirmed clinical diagnosis and severity of AD neuropathology

Boxplot of biomarkers with DX after adjusting for age, sex and time to death

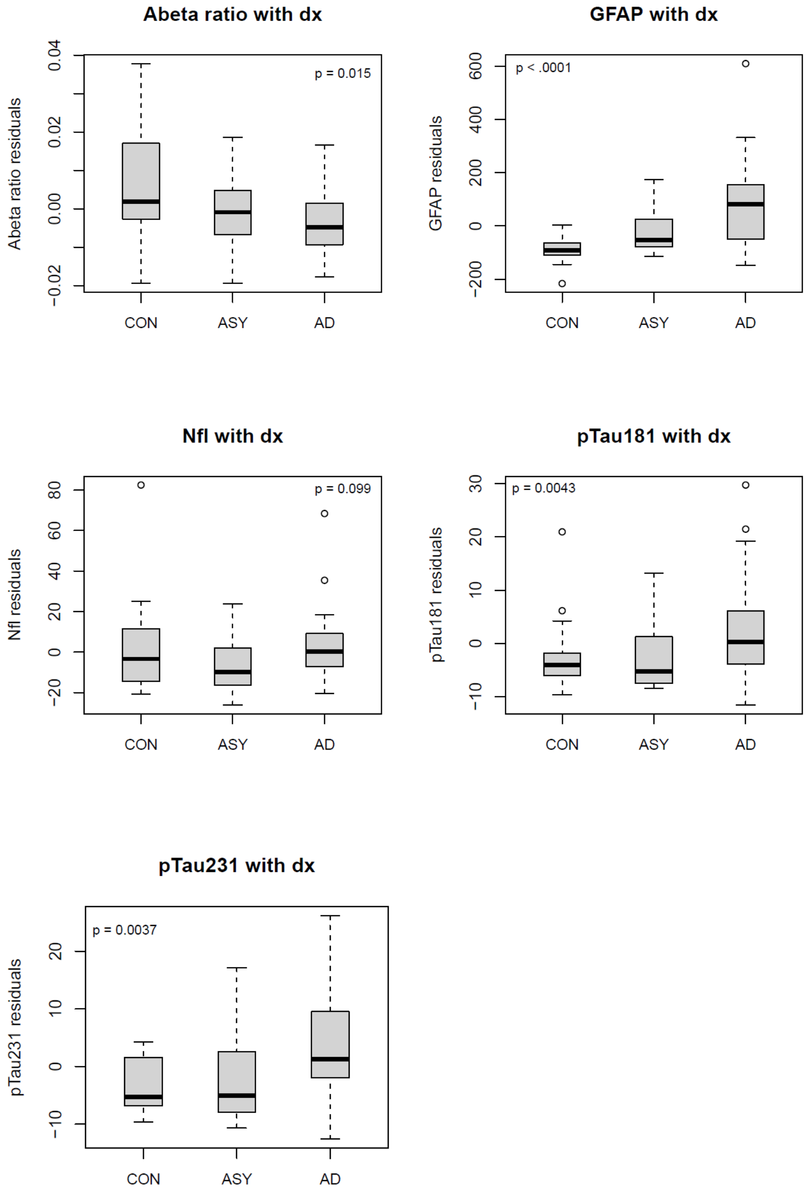

### Boxplot of biomarkers with Braak scores after adjusting for age, sex and time to death

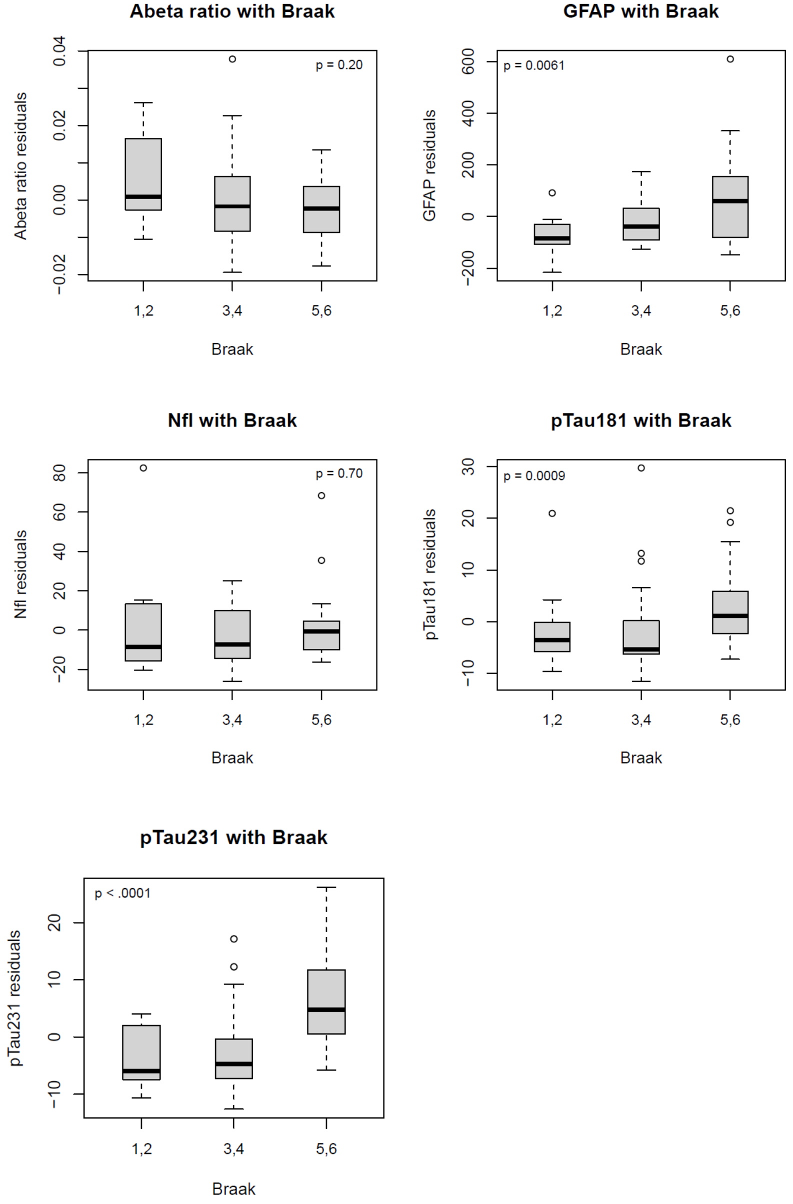

Boxplot of biomarkers with CERAD scores after adjusting for age, sex and time to death

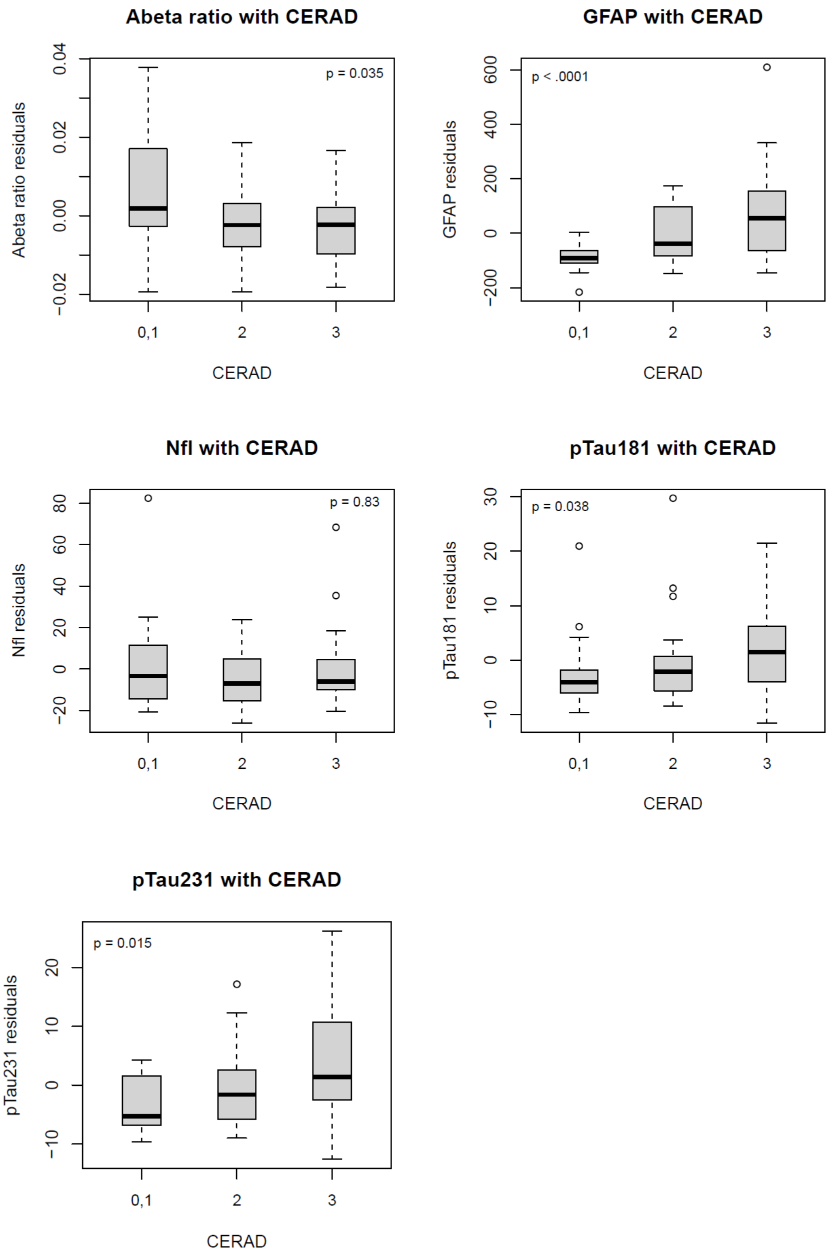


Global p-values (top left or right corner); Diagnosis (dx); Consortium to Establish a Registry for Alzheimer's disease (CERAD); Control (CON); asymptomatic Alzheimer's disease (ASY); Alzheimer's Disease (AD); Glial Fibrillary Acidic Protein (GFAP); neurofilament-light chain (NfL); Beta-Amyloid (Aβ); Aβ-42/Aβ-40 (Aβ ratio); Phosphorylated Tau (p-tau)

**Supplementary Table 6.** Partial spearman correlations between biomarkers and Braak, CERAD scores adjusting for age, sex and time to death:

| | Spearman partial correlation | |
|---|---|---|
| r (P value) | Braak | CERAD |
| Abeta ratio | -0.27<br>p = 0.050 | -0.32<br>p = 0.016 |
| GFAP | 0.43<br>p = 0.0004 | 0.60<br>p < .0001 |
| Nfl | 0.088<br>p = 0.49 | 0.063<br>p = 0.63 |
| pTau181 | 0.43<br>p = 0.0004 | 0.31<br>p = 0.014 |
| pTau231 | 0.58<br>p < .0001 | 0.37<br>p = 0.0034 |

Model covariates included age, sex, race and eGFR.

Consortium to Establish a Registry for Alzheimer's disease (CERAD); Glial Fibrillary Acidic Protein (GFAP); neurofilament-light chain (NfL); Beta-Amyloid (Aβ); Aβ-42/Aβ-40 (Aβ ratio); Phosphorylated Tau (p-tau)

#### Supplementary Table 7. GFAP differences between 5xFAD and WT mice

**Cortex (Brain GFAP levels)**

| | TG - mean (SE) | WT - mean (SE) | difference (p-value) |
|---|---|---|---|
| **3 mo** | 5.125 (1.813) | 5.238 (0.992) | 0.957 |
| **7 mo** | 57.738 (16.586) | 14.313 (6.137) | 0.028 |

| | TG - mean (SE) | WT - mean (SE) | difference (p-value) |
|---|---|---|---|
| **3 mo** | 184.15 (20.141 | 48.788 (3.325) | p<0.001 |
| **7 mo** | 673.7 (74.849) | 68.513 (6.394) | p<0.001 |

**Plasma GFAP**

| | TG - mean (SE) | WT - mean (SE) | difference (p-value) |
|---|---|---|---|
| **3 mo** | 25367.81 (2988.208) | 25262.5 (2817.756) | 0.9797 |
| **7 mo** | 23230.38 (2738.476) | 18748.38 (2416.069) | 0.24 |

**GFAP plasma - GFAP brain cortex**

| | TG - corr (p-value) | WT - corr (p-value) | Combined - corr (p-value) |
|---|---|---|---|
| **3 mo** | -0.3652 (0.3737) | 0.1988 (0.637) | -0.1453 (0.5914) |
| **7 mo** | -0.7457 (0.0337) | -0.1634 (0.699) | -0.0916 (0.7359) |

**GFAP plasma - GFAP brain hippocampus**

| | TG - corr (p-value) | WT - corr (p-value) | Combined - corr (p-value) |
|---|---|---|---|
| **3 mo** | -0.136 (0.7482) | 0.4607 (0.2507) | -0.1381 (0.61) |
| **7 mo** | -0.3912 (0.338) | 0.5163 (0.1902) | 0.3022 (0.2552) |

Glial Fibrillary Acidic Protein (GFAP); months (mo); correlation (corr); standard error (SE)
